# Transcriptome-wide association study identified candidate genes associated with polycystic ovary syndrome

**DOI:** 10.1101/2023.02.09.23285661

**Authors:** Yingzhou Shi, Changrong Li, Hongjie Yu, Xinyu Chen, Ping Shi, Chao Xu

**Author notes:** Corresponding author: Chao Xu. These authors contributed equally to this work (Yingzhou Shi, Changrong Li, Hongjie Yu).

## Abstract

Polycystic ovary syndrome (PCOS) is one of the most common metabolism and endocrine disorder affecting women of reproductive age. However, the genetic mechanisms of PCOS remain largely elusive. A transcriptome-wide association study(TWAS) was conducted to identify the genes associated with PCOS using gene expression references of the whole blood and ovary. Then mRNA expression profiling analysis was performed to identify common genes with TWAS. Genes detected by TWAS were subjected to Gene Ontology(GO), pathway enrichment and protein-protein analysis. TWAS identified 422 genes for the whole blood and 177 genes for ovary, such as EML6 (TWAS.P = 6.33E-05), GRAP (TWAS.P = 0.000211), MAFTRR(TWAS.P = 1.21E-05). Functional enrichment analysis detected biological processes including ncRNA metabolic process, microtubule-related pathways. The present study identified a number of potential PCOS genes and biological processes. These results may provide insight into the genetic basis of PCOS.

## Introduction

Polycystic ovary syndrome (PCOS), characterized by hyperandrogenism, irregular anovulation, and polycystic ovarian changes, is one of the most common reproductive metabolic disorders affecting reproductive-aged women. It is estimated that PCOS affect 6–20% of women^1^, and approximately 80% of those PCOS are infertile^2^. Although studies have suggested that obesity, age at menopause, insulin resistance/hyperinsulinemia, sex hormone-binding globulin (SHBG) levels, and depression may be risk factors for PCOS, its potential etiologies remain uncertain^3^.

Genetic factors are thought to be important in the pathogenesis of PCOS. For example, researchers have found that PCOS is twice as common in monozygotic twins as in dizygotic twins and conclude that genetic factors account for about 79% of the liability of PCOS^4^. The prevalence of PCOS is also higher among the mothers and sisters of affected women, suggesting a phenomenon of familial aggregation^5^. Several genome-wide association studies(GWAS) have identified susceptibility loci for PCOS^6–10^. However, the biological significance of their findings remains largely unclear as close to 90% of GWAS-identified SNPs are in the noncoding region. Expression quantitative trait loci (eQTL) are genomic loci that are associated with gene transcript levels. Gene expression is a critical intermediate phenotype between genes and diseases. Such integration of GWAS and eQTL is effective in identifying candidate genes associated with diseases.

The availability and cost of specimens, as well as intrinsic factors, small effects, make large-scale expression-trait associations challenging. To address these issues, transcriptome-wide association studies (TWAS) were developed, which combine gene expression data with large-scale genome-wide association studies. First, datasets (e.g. GTEx) with matched expression data and genotype were used for building a prediction model. Then GWAS data are combined with these models to assess the association between predicted gene expression and disease risk. A growing number of researchers are using TWASs to identify disease-associated genetic loci^11–13^.

In the present study, we conduct TWAS and mRNA expression profiling analysis to identify genes that are associated with PCOS. Initially, TWAS was performed to identify genes associated with PCOS risk. Then mRNA expression profiling analysis was carried out to identify common genes between TWAS. For the genes identified by TWAS, we conducted an analysis of gene ontology and pathway enrichment to explore their biological functions and related pathways.

## Method

### GWAS summary data for PCOS

We obtained PCOS GWAS summary data from a GWAS meta-analysis of seven cohort studies that included 10074 patients with PCOS and 103164 controls of European ancestry^7^. The cohort studies include Rotterdam(cases: 1184, controls:5799), UK (cases: 670,controls: 1379), EGCUT(cases:157,controls 2807), deCODE(cases: 658,controls: 6774), Chicago (cases: 984, controls: 2963), Boston(cases: 485,controls: 407), 23andMe(cases: 5184, controls: 82759). Diagnosis of PCOS was made based on National Institutes of Health criteria^14^, Rotterdam criteria^15^, or self-report.

### TWAS analysis of PCOS

TWAS analysis was conducted using FUSION software. Pre-computed gene expression weights combined with PCOS summary statistics were used to calculate the association between single gene and PCOS. We derived the gene expression weights for whole blood and the ovary from the FUSION website (http://gusevlab.org/projects/fusion). In FUSION, expression weights were calculated using five linear models, including the best linear unbiased prediction model (BLUP), Bayesian sparse linear mixed model (BSLMM), least absolute shrinkage and selection operator (LASSO), Elastic Net, and top single nucleotidepolymorphisms (SNPs). In the present study, a TWAS *P*-value was calculated for each gene within the whole blood and ovary for European populations.

### mRNA Expression profiles of PCOS

A genome-wide mRNA expression profile of PCOS was used to identify differentially expressed genes (DEGs). The PCOS expression data were downloaded from the Gene Expression Omnibus (GEO) Datasets GSE34526 (https://www.ncbi.nlm.nih.gov/geo/query/acc.cgi?acc=GSE34526)^16^. Gonadotropin-stimulated granulosa cells were obtained from 7 patients with PCOS and 3 healthy individuals. GEO2R was used to screen for differentially expressed genes^17^. Genes with a folding change (FC) >2 or <0.5 (|log2FC |> 1) and p-value < 0.01 were considered to be differentially expressed genes.

### Functional Enrichment

Metascape (https://metascape.org/gp/index.html) was used to perform gene ontology and pathway enrichment analyses of the genes identified by TWAS. Metascape analysis is based on databases, including Gene Ontology, KEGG, Reactome Gene Sets, and WikiPathways. A hypergeometric test is used to identify enhanced ontology terms, and the p-value is corrected for false discovery rates. Molecular Complex Detection (MCODE) algorithm was used to extract densely connected regions^18^.

## Results

### TWAS results of PCOS

A total of genes were identified by TWAS in blood and ovary tissue.

The TWAS identified 422 genes with a p value < 0.05 for the whole blood, such as EML6 (TWAS.P = 6.33E-05), GRAP (TWAS.P = 0.000211), PTGER2(TWAS.P= 0.000403)(Supplementary Table1 and Table2). For the ovary tissue, TWAS identified 177 genes with a p value < 0.05, such as MAFTRR(TWAS.P = 1.21E-05), NEIL2 (TWAS.P = 3.22E-05), ZNF555(TWAS.P= 0.000364). 52 genes with p value < 0.05 were detected in both whole blood and ovary TWAS analysis (Supplementary Table3). Table 1 and Figure1 show the top 20 genes associated with PCOS as determined by TWAS.

**Table 1.** Top 20 genes identified by TWAS analysis

| Gene | Ensemble | Best GWAS ID | TWAS P-value | Tissue |
| --- | --- | --- | --- | --- |
| MAFTRR | ENSG00000261390 | rs17767383 | 0.0000121 | Ovary |
| NEIL2 | ENSG00000154328 | rs2686206 | 0.0000322 | Ovary |
| EML6 | ENSG00000214595 | rs11898747 | 0.0000633 | Whole blood |
| GRAP | ENSG00000154016 | rs11652784 | 0.000211 | Whole blood |
| ZNF555 | ENSG00000186300 | rs887500 | 0.000364 | Ovary |
| RPS26 | ENSG00000197728 | rs3741499 | 0.000378 | Ovary |
| PTGER2 | ENSG00000125384 | rs781367 | 0.000403 | Whole blood |
| RPS26 | ENSG00000197728 | rs3741499 | 0.000428 | Whole blood |
| MAPKAPK5-AS1 | ENSG00000234608 | rs6489819 | 0.000465 | Whole blood |
| UBE2Q2L | ENSG00000259511 | rs28600121 | 0.000507 | Ovary |
| ZNF556 | ENSG00000186300 | rs887500 | 0.000532 | Ovary |
| ARMH2 | ENSG00000260286 | rs9393592 | 0.000589 | Whole blood |
| RFWD3 | ENSG00000168411 | rs4072222 | 0.000731 | Whole blood |
| MAST3 | ENSG00000099308 | rs11666281 | 0.000758 | Whole blood |
| ICA1 | ENSG00000003147 | rs10952107 | 0.000763 | Whole blood |
| RNF112 | ENSG00000128482 | rs11652784 | 0.000811 | Whole blood |
| SERPINB10 | ENSG00000242550 | rs2162352 | 0.000971 | Whole blood |
| HMGN1 | ENSG00000205581 | rs7279465 | 0.00113 | Whole blood |
| ARHGAP1 | ENSG00000175220 | rs6485690 | 0.00122 | Ovary |
| SLC41A1 | ENSG00000133065 | rs6681714 | 0.00126 | Whole blood |

**Figure1.**
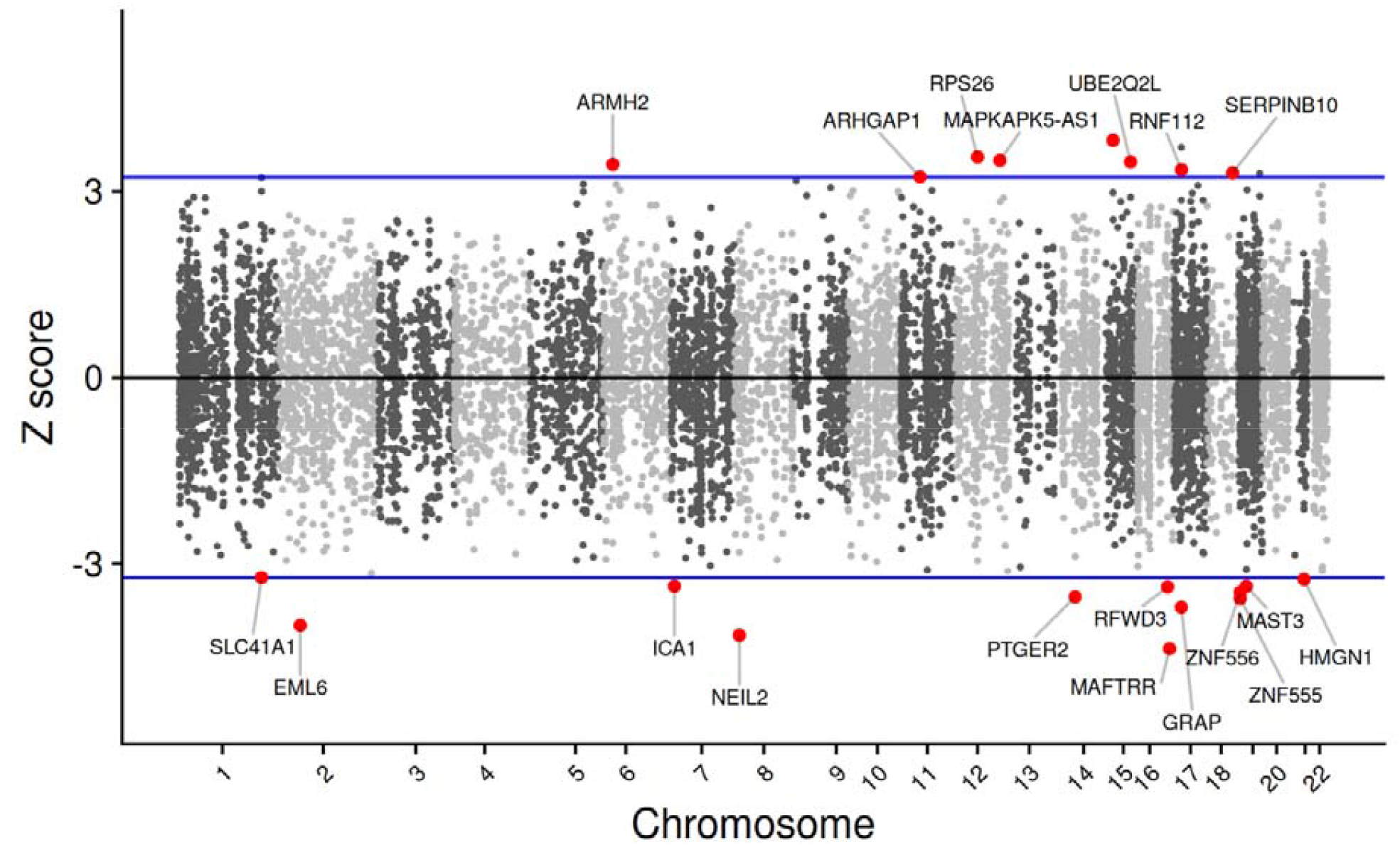
Manhattan plot of TWAS-identified top20 genes associated with PCOS The x-axis represents the physical location of each gene, while the y-axis represents the Z score of association between that gene and PCOS.

### Genes shared between TWAS and mRNA Expression Profiling

The TWAS analysis and the mRNA expression analysis detected 18 common genes, such as CAMK1D, COLEC12 and PLCB2.(Table 2). A Venn diagram of TWAS versus common genes identified by mRNA expression profiling are shown in Figure2.

**Table 2.** Common gene identified by TWAS and mRNA expression profiling

| Gene symbol | TWAS.Z | TWAS.P | Log2fold change | P.value |
| --- | --- | --- | --- | --- |
| CYBRD1 | -2.29174 | 0.02192 | 3.3603695 | 0.00470417 |
| RAB31 | -2.2727 | 0.023 | 1.6377338 | 0.00929241 |
| NAGA | 2.0323 | 0.04213 | 1.8187126 | 0.0084649 |
| CAMK1D | 3.01 | 0.00262 | 2.0792639 | 0.00507069 |
| NUCKS1 | 3 | 0.0027 | 1.505372 | 0.00697271 |
| PLCB2 | 2.98094 | 0.002874 | 2.3351678 | 0.00599101 |
| COLEC12 | 2.7632 | 0.005724 | 1.4609694 | 0.00460956 |
| TTC27 | 2.327847 | 0.0199 | -1.1913658 | 0.00690472 |
| GTF2IRD2 | -2.28205 | 0.022486 | 3.3187845 | 0.00244768 |
| LIMD2 | -2.25126 | 0.024369 | 2.8804695 | 0.00075794 |
| MS4A6A | -2.173 | 0.02978 | 1.8833963 | 0.00947628 |
| PDE8B | 2.15237 | 0.03137 | -2.0226354 | 0.00865061 |
| LIME1 | 2.151515 | 0.03144 | 1.2175341 | 0.0096489 |
| RNASE2 | 2.0625 | 0.03916 | 2.7763519 | 0.00248818 |
| LOC100505716 | 2.03598 | 0.0418 | -2.1059969 | 0.00336991 |
| KCNJ15 | 1.99801 | 0.04572 | 2.3330272 | 0.00287839 |
| TGFBI | -1.97181 | 0.04863 | 3.2674614 | 0.00011652 |
| CARMIL1 | -1.9693 | 0.048924 | -2.5048194 | 0.00130639 |

**Figure2.**
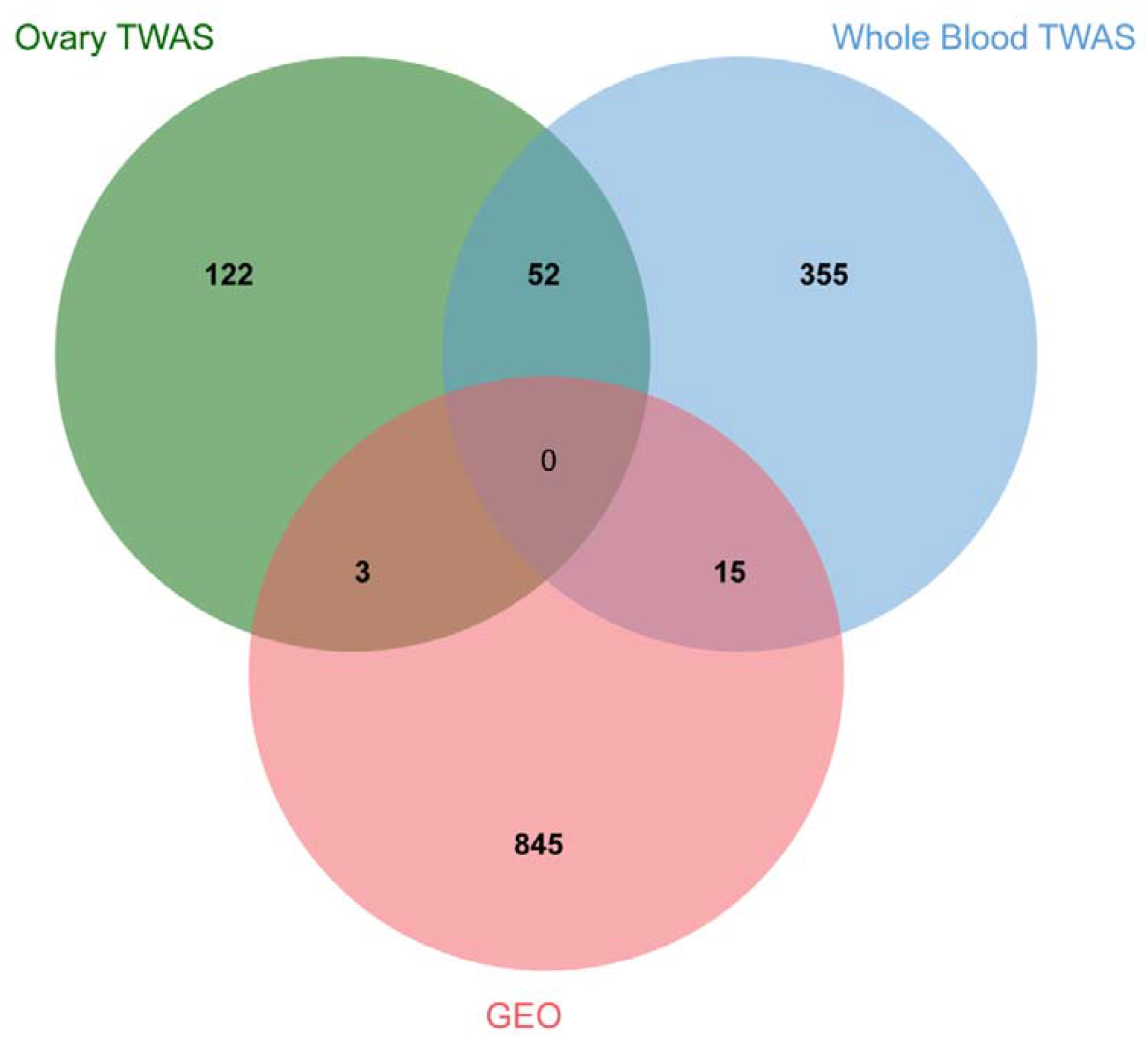
Venn diagram of gene identified by TWAS and mRNA expression profiling

### Gene Enrichment Analysis of genes identified by TWAS

In order to obtain an overall impression of the characteristics of these genes identified by TWAS, a total 547 genes were submitted to Metascape performing GO and pathway enrichment, and protein-protein interaction analysis.

GO and pathway enrichment analysis showed that TWAS-identified genes were enriched in ncRNA metabolic process, rRNA metabolic process, transport along microtubule and so on(Figure 3). MCODE enrichment analysis based on PPI enrichment for TWAS-identified genes showed 11 PPI modules(Figure 4). The top enrichment term of MCODE1 included 13 proteins distrusted in three major clusters: “Metabolism of RNA”, “Major pathway of rRNA processing in the nucleolus and cytosol”, “rRNA processing in the nucleus and cytosol”. MCODE2, which consists of 7 proteins was mostly enriched in “Interleukin-20 family signaling”, “JAK-STAT signaling pathway”, “Regulatory circuits of the STAT3 signaling pathway”.

**Figure 3.**
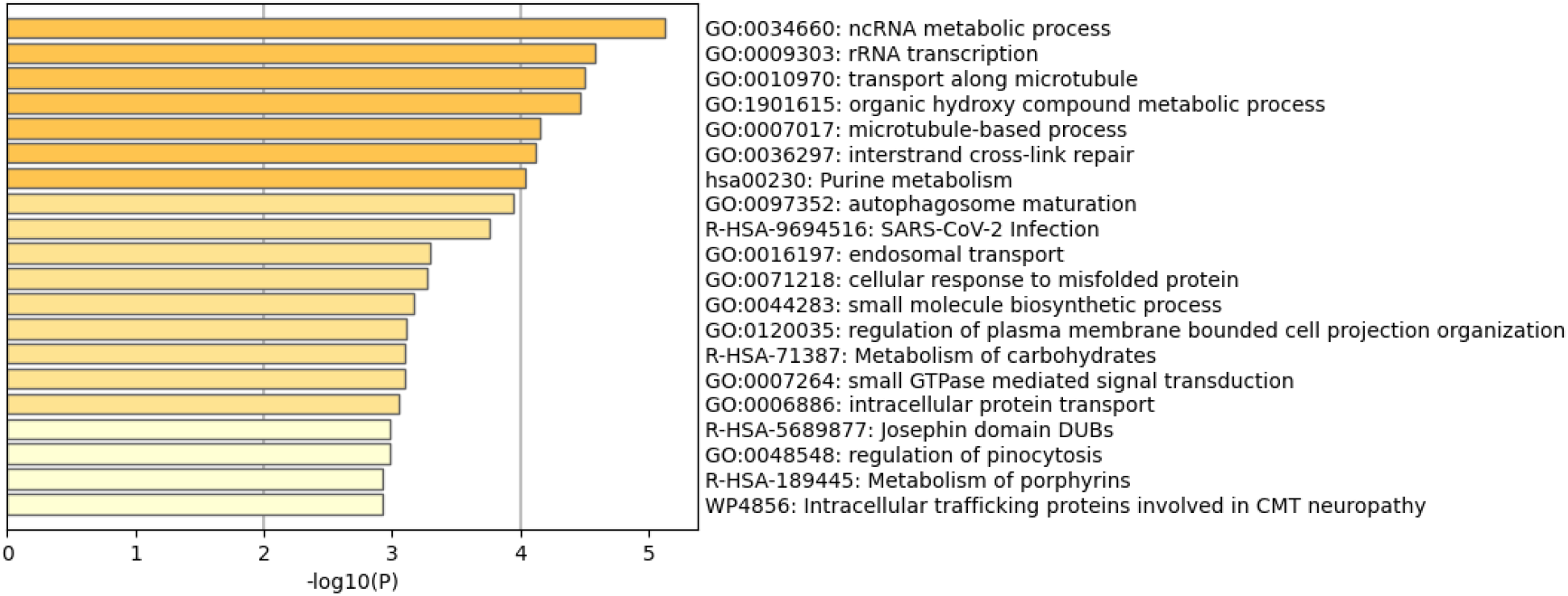
GO enrichment analysis of the genes identified by TWAS

**Figure 4.**
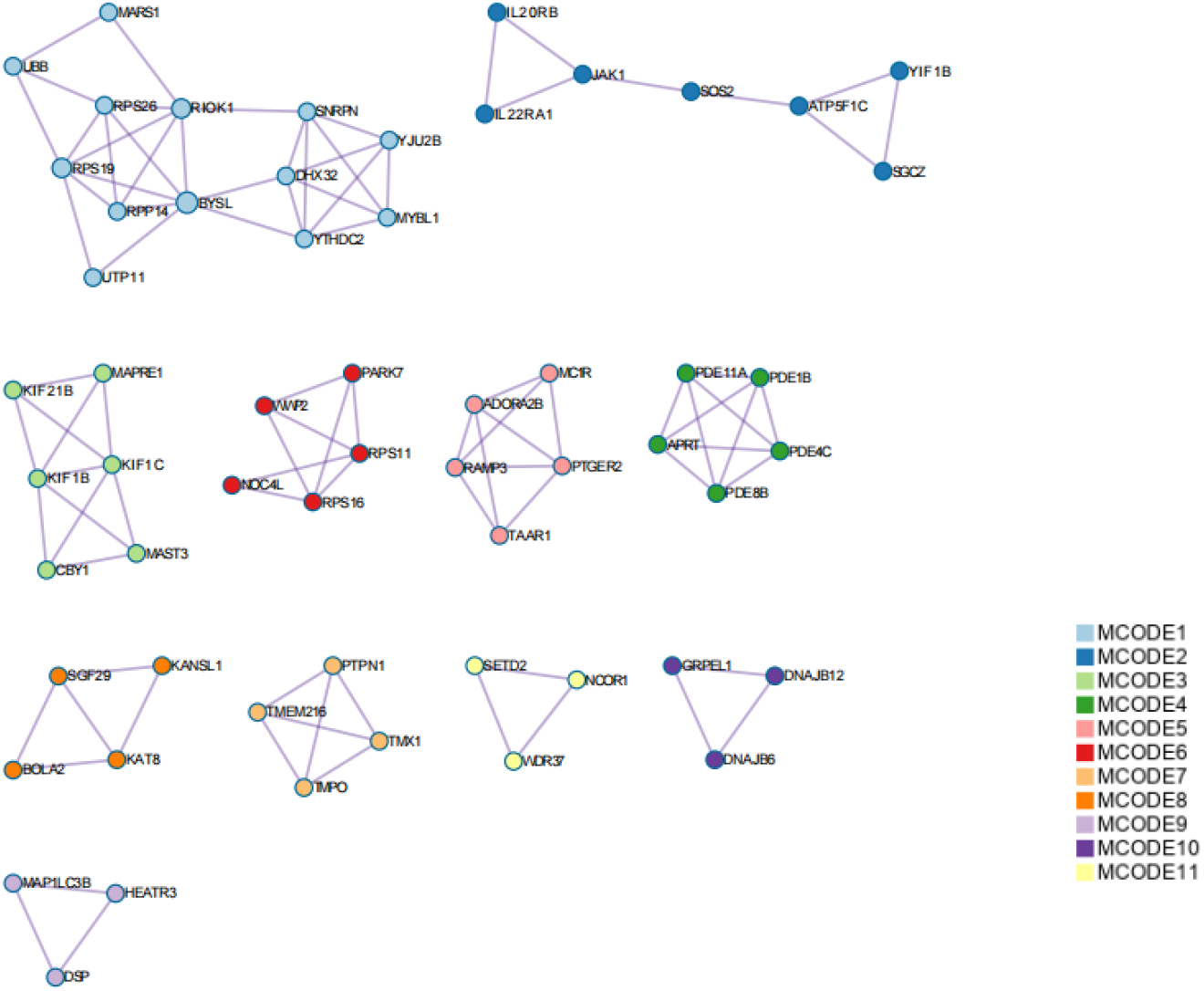
Protein-protein interaction network of genes identified by TWAS

## Discussion

The TWAS method is a powerful tool that utilizes genetic variation and gene expression to identify genes whose cis-regulated expression is associated with complex traits. In present study, we performed TWAS to detect candidate genes closely associated with PCOS. Further integrative analysis of TWAS and mRNA expression profiling identified 18 common genes. To the best of our knowledge, this is the first TWAS integrated with mRNA expression profiling analysis for PCOS. The results of our study provide new insight into the genetic mechanism behind PCOS. Our TWAS identified several genes that have been previously reported in GWAS and post-GWAS, such as RPS26^19^, NEIL2^8^, and ZNF556^19^ as well as novel genes that have never been reported.

NEIL2 is is the gene encoding a member of the Fpg / Nei DNA glycosylase family. The encoded enzyme is involved in DNA repair mainly by cleaving oxidatively damaged bases and introducing DNA strand breaks through its base cleavage enzyme activity.

The NEIL2 gene encodes NEI endonuclease VIII-like 2, one of the DNA glycosylases involved in DNA repair^20^. DNA damage has been demonstrated to play a role in the pathogenesis of PCOS^21^. Further experiments are needed to determine the relationship between NEIL2 and PCOS.

EML6 is the only member of the EML family that is preferentially expressed by oocytes in the mouse ovary^24^. There is a colocalization of EML6 with meiotic spindles, and this specialized localization plays a critical role in maintaining spindle integrity and homologous chromosome segregation.

RPS26, located on 12q13.2, is a gene encoding a ribosomal protein, which is a component of the 40S subunit and belongs to the ribosomal protein S26E family^22^. The knockdown of Rps26 in mouse oocytes has previously been shown to result in delayed oocyte development and even the cessation of chromatin structural transition in oocytes, resulting in premature ovarian failure (POF)^23^.

GO and pathway enrichment indicated that ncRNA metabolic and microtubule-related pathways were enriched. The non-coding RNAs (ncRNAs) are functional RNAs that are not coded for protein production but serve as important regulators of a wide range of biological events^25^. Several studies have indicated that ncRNAs play a role in the occurrence and development of PCOS. The expression of sncRNAs was significantly different in serum, granulosa cells (GCs), follicular fluid (FF), and other tissues between PCOS patients and the general population^26,27^. Accordingly, our results further confirm that ncRNA may play a crucial role in the development of PCOS.

Microtubules, which are polymers containing α- and β-tubulin heterodimers, make up the oocyte spindle^28^. An abnormal spindle can affect the development, differentiation, and growth of a germ cell directly. Recent research has identified several mutations of spindle-related proteins that lead to infertility. An example of this is the TUBB8 gene, which encodes an isoform of beta-tubulin that is specific to humans and primates. TUBB8 mutations in women lead to a range of spindle defects during oocyte meiosis^29^. The enriched microtubule-related pathways may suggest that microtubules contribute to infertility in women with PCOS.

## Conclusion

In summary, this TWAS study identified novel and susceptible genes and potential pathways for PCOS. Further studies are needed to explore the potential mechanism.

## Data Availability

https://www.repository.cam.ac.uk/handle/1810/283491

## Acknowledgements

We would like to thank the participants and the investigators for generating the publicly available dataset. The scientific calculations in this paper have been done on the HPC Cloud Platform of Shandong University.

